# “What matters in help-seeking and disclosure intent of intimate partner violence during the COVID-19 pandemic: Similarities and differences across demographic groups”

**DOI:** 10.1101/2025.05.23.25328223

**Authors:** Christina Palantza, Maxine Davis, Anke B. Witteveen, Diana M. Padilla Medina

## Abstract

The COVID-19 pandemic increased Intimate Partner Violence (IPV) rates internationally and disrupted health services. Physical health concerns were prominent due to the direct impact of IPV. However, beyond physical injuries the pandemic also exacerbated other risk factors linked to IPV, such as deteriorating mental health, which can further hinder help-seeking for IPV. As access to health care services became more restricted, those experiencing IPV faced even greater barriers to help-seeking. No study has examined the factors related to IPV help-seeking intent during the earliest stage of the pandemic; therefore, the aim of this study was: To examine the comparative impact of the COVID-19 burden and health factors on IPV help-seeking and disclosure intent using quantitative data. A cross-sectional survey, named The Health and IPV Study (HIPVS) was conducted in the USA in April 2020. Among other health-related constructs, HIPVS assessed historical, recent, and current health status (mental and physical), IPV (victimization and perpetration), IPV help-seeking and disclosure intent. Subgroup analyses were done by gender, age and income level. Generalized Linear Mixed Effects models were used (N=1346). Number of COVID-19 cases and PTSD symptomology predicted higher help-seeking intent, but experience of violence correlated with lower disclosure intent. There were significant differences among demographic groups and inconsistency in the reporting of violence was a notable issue. The findings on mental health support the knowledge in existing literature. The most important limitation was the phrasing of the questions, particularly those which focused on intent instead of behavior. Mental and medical healthcare providers should prioritize IPV screening. Access to care should be maximized through continued improvement/expansion of online service provision and changes in healthcare policy that remove barriers (such as lapse in medical insurance coverage or financial burden).

## INTRODUCTION

Intimate Partner Violence (IPV) is any form of aggression or violence towards one’s intimate partner (1) and a major public health issue across the globe (2). The COVID-19 pandemic, which started affecting a great deal of the world in March 2020, has repeatedly been shown to increase the prevalence, severity and frequency of IPV internationally (3). The evidence regarding prevalence initially appeared to be mixed, with some studies reporting decreases (4), and other studies suggesting increases (5, 6). These differences are now explained by help-seeking bias, meaning that reduced access to services was a major reason for fewer reports in many settings (7). To further complicate the matter, discrepancies have been found in the reports of survivors and providers (7).

Help-seeking has had a pivotal role in establishing the state of the matters in IPV during the context of the pandemic. The demand for services was high but not always met (8) (9) with IPV worsening and barriers to help-seeking rising (10). Access to IPV services worsened during the pandemic (11, 12) as it would be expected, given the lockdowns or social distancing measures, and the overloading of health systems. At the same time, providing health and social care delivery became more challenging (8, 13). Consequently, the help-seeking patterns for IPV changed throughout the course of the pandemic (14), but correlates to such changes have not yet been investigated (15).

Formal help-seeking is necessarily linked with disclosure of IPV to professionals. The most common barriers to disclosure are fear of retaliation by the partner (16–18) and feeling trapped or dependent on the partner (19). For many, the stay-at-home and shelter-in-place orders created an environment which increased such feelings. On top of that, literature spanning decades indicate that IPV-affected individuals are much more comfortable and likely to disclose IPV, when there is a personal, compassionate and understanding approach by the provider (20). Such an approach is less likely to be adopted by health-care professionals in the context of widespread disaster (i.e. the pandemic), while providers face numerous challenges/uncertainty in the workplace (13), experience increased private life stressors, and heavier workloads. Moreover, survivors tend not to communicate with providers about IPV, when they believe there is a lack of time (21), which was a reality throughout the pandemic—especially during the earliest stage, and studies have shown that there were missed opportunities for IPV screening (22). Survivors are more likely to disclose IPV when screened for it, rather than to mention it themselves to the providers voluntarily without probe (23, 24). However, the encouraging and supportive approach is *necessary* for the screening to be acceptable (25).

To more efficiently manage the rising need for services in their limited availability, in the context of the COVID-19 pandemic crisis, it would have been imperative to pinpoint which factors impacted help-seeking (26), the prerequisites of disclosure. There is already some evidence that help-seeking did not actually decrease during the pandemic, rather referrals decreased due to practical constraints (27). When screened by healthcare providers disclosures were low compared to the actual prevalence (28). This objective for greater understanding is pressing, as climate change is speculated to incur more crises, including pandemics (29).

The COVID-19 pandemic has had widespread effects on public health, with mental health emerging as a particularly vulnerable area (30). Poor mental health and substance abuse are among the most common and strongest correlates of IPV. IPV has been shown to take a heavy toll on survivors’ mental health (31, 32), but perpetrators very often suffer from mental disorders and abuse substances too (33, 34). Substance abuse, especially of alcohol, increased in the context of the pandemic (35), and this has been directly connected to increased IPV risk (36, 37). Mental health issues play a significant role in help-seeking behaviors as well (38), with some studies identifying mental health issues as a barrier to help-seeking (39), while other studies describe experiencing poor mental health as motivation to seek help (40, 41).

Improving mental health may be seen as second to creating a need for services when compared to the violence itself (42). Severity of violence and experiencing multiple types of violence (psychological, physical, sexual) at once, have been consistently shown to lead to help-seeking (43) (44–47). Therefore, descriptions of violence need to be considered when assessing factors influencing help-seeking.

Apart from mental health and violence, there is an instrumental role of structural and practical factors related to help-seeking. During the COVID-19 pandemic, these systemic barriers—such as restricted mobility, service availability, and financial instability—may have had a greater impact than interpersonal factors. Evaluating the relative influence of these barriers, alongside mental health and violence severity, is essential for a comprehensive understanding of IPV-related help-seeking.

In addition to intrapersonal and external factors, the public’s top concern during the COVID-19 pandemic was physical health. Physical health is well known to be particularly compromised among survivors of IPV (45, 48). People with severe or chronic physical health issues would presumably prioritize these conditions instead of IPV during the pandemic for several reasons. For example, being reluctant to visit services out of fear of getting infected with COVID-19, or avoiding leaving the house, even when allowed by social distancing measures. With that in mind, we hypothesized that poor physical health was associated with decreased help-seeking for IPV during the pandemic.

Help-seeking in the pandemic has been researched mostly through the records of health services, the police, and helplines (14, 27, 49–51). While it is acknowledged that there has been a significant gap between the need and access to services (7), survivors have expressed perceiving services as harder to access (52), especially those belonging to previously disadvantaged and vulnerable groups (53). In order to capture the full scope of help-seeking behavior in the pandemic and the factors that influence it, it is necessary to study the intent to seek help and the intent to disclose IPV among individuals affected by IPV, and not just service records.

Based on the existing evidence, the primary research question of this study is: What is the comparative impact of the state-level COVID-19 burden, mental and physical health, alcohol abuse, PTSD and IPV on help-seeking and disclosure intent? In order to investigate potential demographic differences in further detail, the following research sub-questions were examined:

RQ 1.1: What predicted intention to seek help for IPV?

RQ 1.2: Did correlates of intention to seek help for IPV differ by gender?

RQ 1.3: Did correlates of intention to seek help for IPV differ by age?

RQ 1.4: Did correlates of intention to seek help for IPV differ by income level?

RQ 1.5: Did correlates of intention to seek help for IPV differ by ethnicity?

RQ 2.1: What predicted intention to disclose IPV?

RQ 2.2: Did correlates of intention to disclose IPV differ by gender?

RQ 2.3: Did correlates of intention to disclose IPV differ by age (life-stage)?

RQ 2.4: Did correlates of intention to disclose IPV differ by income level?

RQ 2.5: Did correlates of intention to disclose IPV differ by ethnicity?

## METHOD

### Design, setting and ethics

This study is a secondary analysis of a subset of the sample of a cross-sectional survey which was carried out online among the general population in the United States from April 15th to May 1st 2020, using the Qualtrics platform. The main description of the data collection methodology can be found in (54). The participants gave written informed consent through Qualtrics. The original survey was ethically approved by the Institutional Review Board (IRB) of the University of Texas at Arlington (Protocol number 2020-0193).

### Participants

The convenience sample was comprised of individuals who have signed up to the Qualtrics platform for any survey, driven primarily by the financial rewards of the surveys. Quota sampling by demographics was used to achieve a balanced and representative sample of the US general population. Recruitment for survey participation was described in generic terms to decrease the chance that IPV-involved individuals would increasingly approach or avoid the survey. IPV prevalence data in the entire sample was consistent with prior nationally collected data in the U.S. Only participants that had perpetrated IPV or were victimized by IPV were included in the present study. That meant answering positively to at least one question on psychological, physical, or sexual victimization and/or perpetration.

### Data collection and management

All participants completed the questionnaires online. Responses that were duplicate, completed suspiciously fast or violating the quota were removed (Ν=1750 out of 3750). The data were anonymized. A data sharing agreement was signed between the first authors of the parent study and the present study, and the data were transferred and stored securely through a General Data Protection Regulation compliant platform. The data for this analysis were first accessed on March 2nd, 2022.

### Operationalization of variables

#### Independent variables

The independent variables were physical health, mental health, alcohol abuse, post-traumatic stress disorder (PTSD) symptoms, COVID-19 burden in the state, changes in daily life due to the pandemic, psychological victimization, perpetration of psychological violence, physical victimization, perpetration of physical violence, sexual victimization, perpetration of sexual violence, and bidirectional violence.

The classification of physical health status from the American Society of Anesthesiologists (55) was adapted to classify health status. The questionnaire included a list of several physical conditions and health symptoms. The participants could answer if they had them currently, in the past, or never. The final classification was: “1: healthy”= having fewer than five of the symptoms listed, “2: mild disease”=having a mild disease, such as an allergy or more than five of the symptoms listed, “3: severe yet not incapacitating disease”=having a serious disease that is not constantly life threatening or incapacitating, such as an autoimmune disease, or diabetes, “4: current constantly life threatening disease”=cancer, and “5: history of serious disease”=checking off any serious disease but only in the past.

Overall mental health was assessed with the shortened version of the Brief Symptom Inventory (BSI-18) (56). Each item is scored 0-3, so the range of scores is 0-72. It has been found to have adequate psychometric properties (56). Since PTSD is one of the most common disorders among people involved in IPV (31), it was included as a separate variable and measured with the International Trauma Questionnaire (ITQ) (57). It consists of six items, representing the three clusters of PTSD symptoms (reexperiencing, avoidance and sense of threat). The items are scored 0-4, and the total score was used. It has high reliability in the US population (57). The survey included questions on mental disorder diagnoses, in the same fashion as the physical ones, and a variable indicating having any mental diagnosis was also used, because individuals who have received a diagnosis often face increased barriers in accessing healthcare services (58, 59). Alcohol abuse was one of the conditions listed, so the participants self-reported whether they had it currently, in the past or never.

The burden of COVID-19 in the state was operationalized as the number of cases in the state on May 1 2020, in the same manner as in Davis and colleagues (54) [i.e. an ordinal variable of four levels (0-4999 cases/5000-9999/10000-23999/24000+)].

Subjective change in one’s daily life might implicitly affect their mental health and decisions regarding help-seeking, so an ordinal variable on changes in daily life was included, based on dichotomous questions about changes experienced. The levels were: 0=no changes, 1=small changes, such changes in social life and starting to work from home, 2=severe changes, such as having lost one’s job or changes in family life.

Perpetration and victimization of IPV were assessed with three different scales, as the survey’s original purpose was to assess the psychometric properties of a newly developed IPV tool. The three scales were the 5-item Extended-Hurt/Insult/Threaten/Scream (Modified E-HITS) (60), the Jellinek inventory for assessing partner violence (J-IPV) (61), and the Revised Conflict Tactics Scale (CTS-2) (62). All of these tools have shown satisfactory psychometric properties; the E-HITS has been identified as an accurate and acceptable screener (60); the J-IPV is a very low-burden tool, as it consists only of 4 items and it is a very robust screener (63). The CTS-2 is more elaborate and serves as a reference. Nevertheless, the participants of the present study often gave inconsistent answers across IPV scales in the three measures. This has been attributed to gender and age disparities in phrasing (64), but assuming that some participants could be ambivalent about reporting, a positive answer in any scale was deemed as sufficient. Further work on the reporting inconsistencies is under preparation. All these scales include questions both on victimization and perpetration.

Psychological violence was coded as a continuous variable, because psychological violence has been shown to have a variety of nuances with different impact on the survivors (65). The coding was done in scores: 0=no violence, 1=only yelling, 2=only insults or only threats, 3=insults and yelling, or threats and yelling, or insults and threats. Past psychological victimization and perpetration were separate variables, as history of psychological abuse can have a lasting effect on mental health (66). Physical and sexual violence were both coded dichotomously, either in absence or presence of any violence. A dichotomous variable on bidirectional violence (i.e. identifying both as a survivor and a perpetrator) was also used, because people affected by bidirectional violence have been found to have varying help-seeking trajectories (67).

#### Outcome variables

Help-seeking intent was measured with one item “If a healthcare provider, such as your primary care provider, were to ask you such questions, to what extent would you be likely to be interested in hearing about resources that may be of help?”, answered with a Likert type scale from “extremely unlikely” to “extremely likely”. Disclosure intent was measured with two items, phrased as: “If a healthcare provider, such as your primary care provider, were to ask you such questions, to what extent would you be likely to discuss these experiences with the provider?”, and “If a healthcare provider, such as your primary care provider, were to ask you such questions, to what extent would you be likely to answer the questions honestly?”. The answering options were the same as help-seeking intent, and the final score was the average of two items. The outcome questions used were corresponding to the E-HITS.

### Statistical analysis

Descriptive statistics of all the variables and Pearson and Spearman correlations between every pair of variables were calculated in SPSS 28 (68). Generalized Linear Mixed Effects Modelling was run in STATA 16 (69). The data were deemed as nested into states, as the intra-cluster correlation coefficient was <.05, after adjusting for unbalanced cluster sizes, using the method proposed by Shan (70). The model was run separately for victimization and perpetration, because the items answered by participants screening positive for perpetration or victimization were coded separately. The model was also run separately for each type of violence, because the types of violence were highly correlated with each other, and there would be an issue of collinearity if they were all entered into the same model. High correlations between the mental health related variables (BSI score, PTSD score, any mental disorder diagnosis, alcohol abuse) would also cause an issue of collinearity in the same model, so the model was run separately with each mental health variable. A sensitivity analysis controlling for changes in daily life was done for every model. The models were also run without any violence variable, to control for any collinearity of the violence with the main variables, as the correlations were high. Lastly, the model was run with each independent variable individually. Two more sensitivity analyses were conducted as well, one considering only the participants who reported violence in the scale of interest for each type of violence (J-IPV for physical violence, and E-HITS for all other types of violence), and one considering only the participants who reported violence on these scales, and their answers were consistent across all scales. The mental health related variables were all tested simultaneously in the models of the sensitivity analyses.

The literature stresses important differences in help-seeking barriers among genders (71, 72) and people with low income (73), in disclosure among sexual minorities (74) and older adults (75). Hence Kruskal-Wallis tests were run for the independent and dependent variables by gender, age, income level, and ethnicity, as well as corresponding subgroup analyses of the models, because the outcomes followed a gamma distribution, skewed towards higher values.

## RESULTS

### Sample descriptives

After excluding those who were not involved in any IPV in the past six months, the final sample consisted of 1346 participants. The total sample of the original study was N=2045, which means that the prevalence of any IPV of any type and severity in the original sample was 65.82%. The final sample of this study was balanced in terms of gender, with 47.3% females, and 2.5% people had a different gender identity or their identity was not represented (detailed descriptives in Table 1). Emerging adults were a fourth of the sample, adults more than a third (35.3%), and older adults 12.8%. Nearly 80% of the sample was heterosexual, and almost 10% had an orientation other than hetero-, homo-, or bisexual (detailed descriptives in Table 1). Regarding ethnicity, 12.9% was African-American, and 13.9% Latinx participants or Caribbean, while 6.5% identified as multi-ethnic. Almost three fourths were employed (73.3%), more than a third had a middle annual income, and a third upper middle income. Three quarters were Christian (44.4% Catholic, 23.1% Protestant, and 10.4% other Christian). More than 90% were born in the USA. The states with the most participants were New York (14.3%) and California (12.3%). Nearly half had faced mild changes in their daily life because of COVID-19, and one quarter had been diagnosed with COVID-19 or someone in their close environment had been infected. A little more than 40% had had a mental disorder diagnosis in their lifetime, and less than a third was classified as healthy. Approximately 90% identified both as a survivor and a perpetrator. Fourteen percent had lifetime alcohol abuse. Past six month physical perpetration and victimization were between 40 and 44% respectively, and sexual perpetration and victimization between 31 and 34% respectively. More details can be found in Table 1. In the following sections with the results of the regression models testing correlates of the outcomes, we only mention the correlates that were confirmed in the majority of the models. The full set of variables that reached significance are presented in the tables.

**Table 1.**
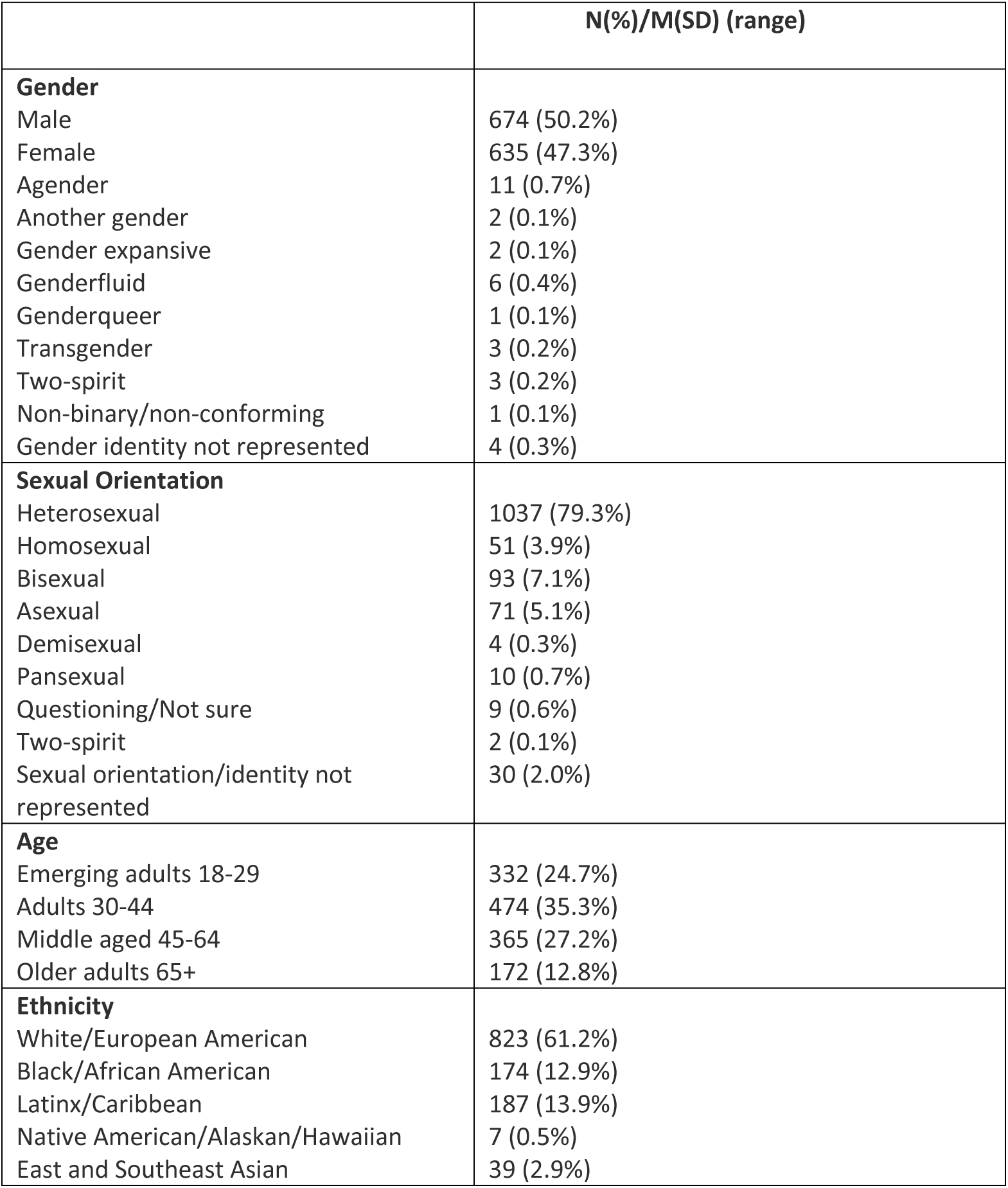
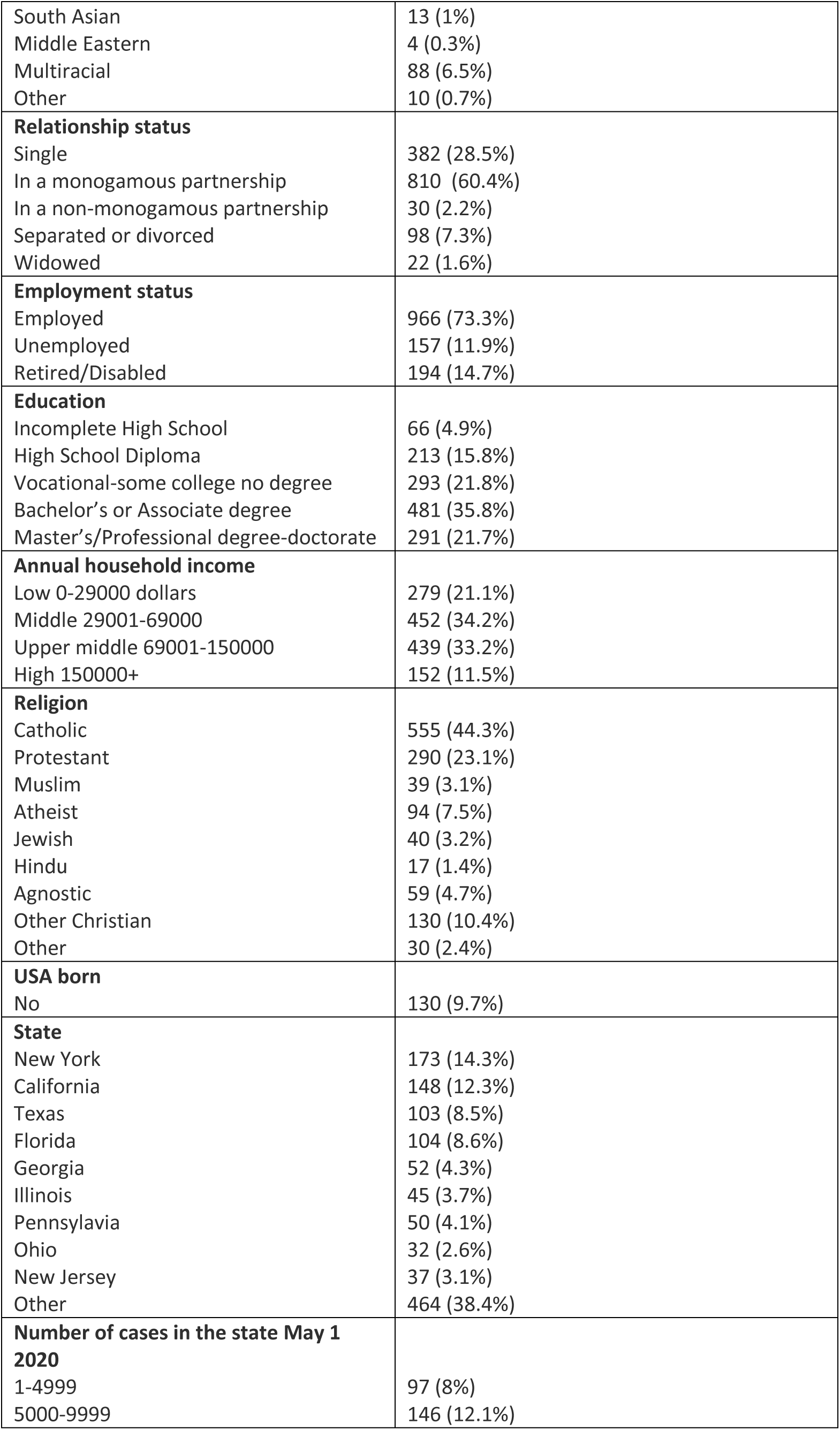
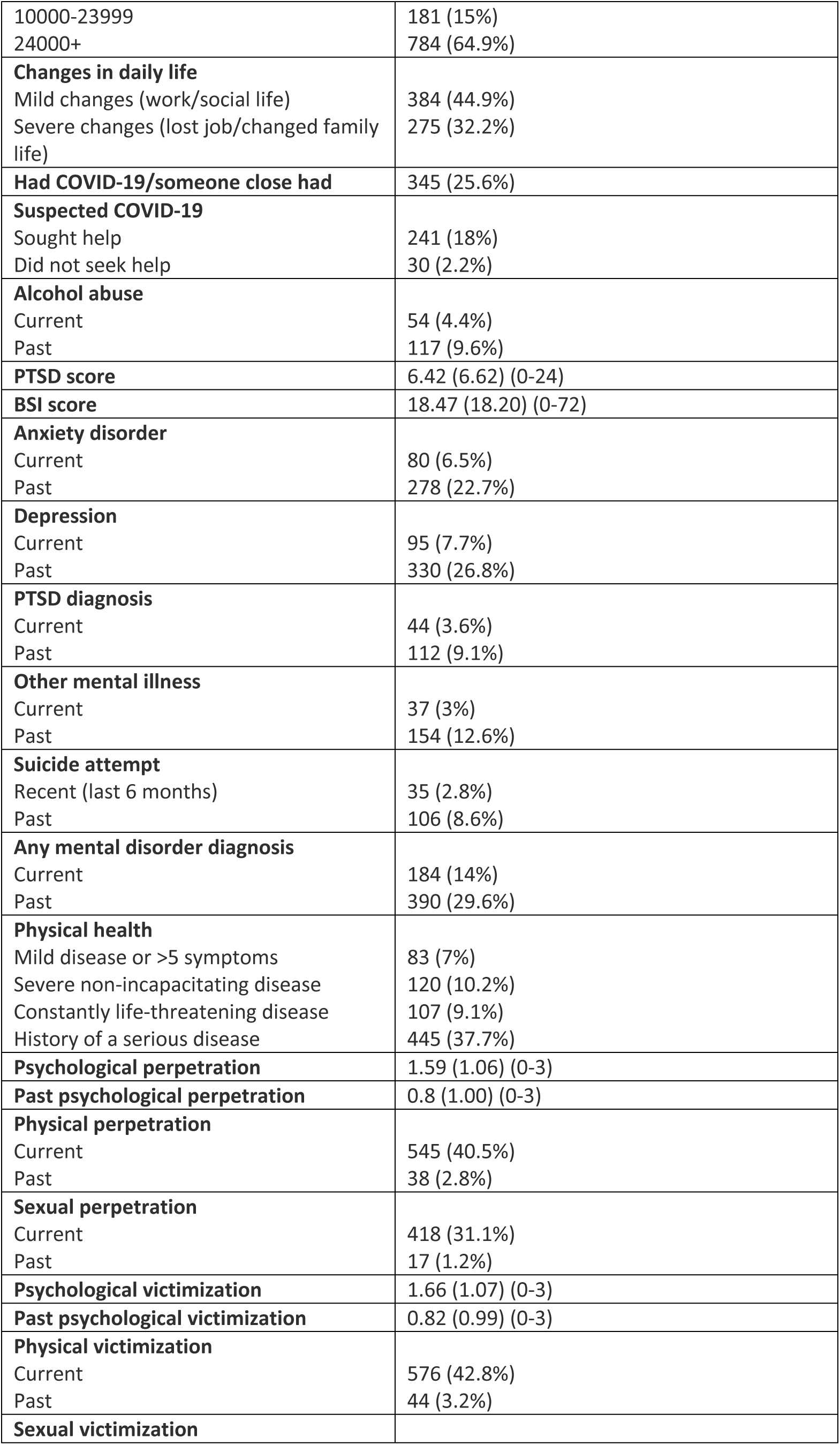
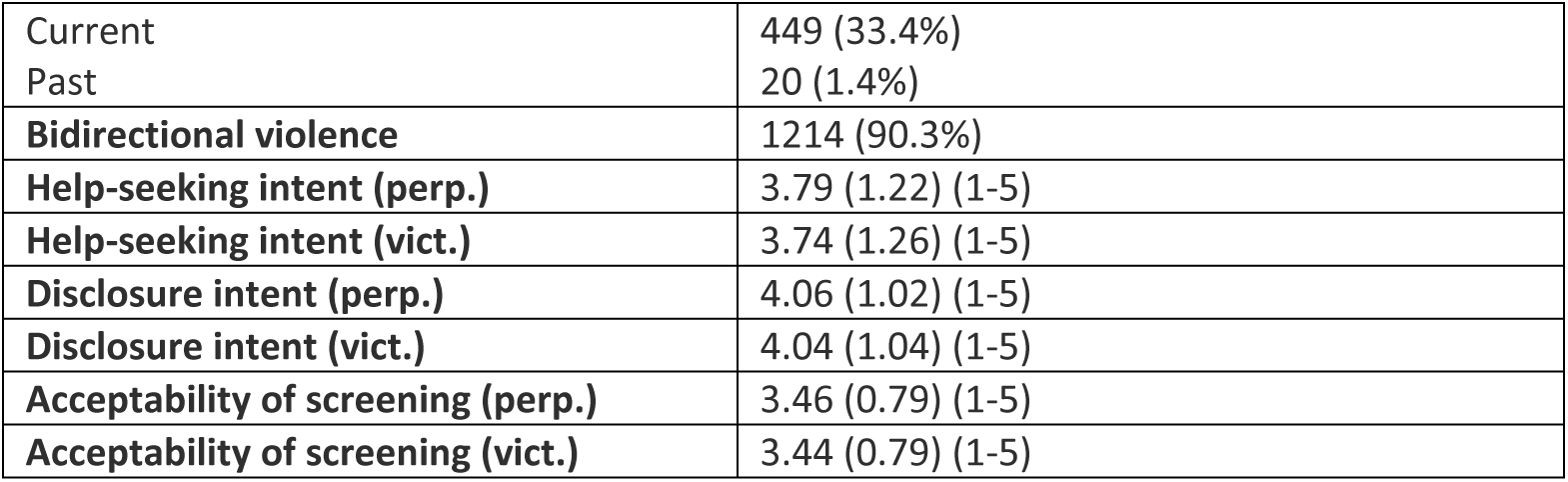
Descriptives of the sample N=1346.

### Help-seeking intent (Table 2)

#### RQ 1.1: What predicted intention to seek help for IPV?

Number of COVID-19 cases in the state and PTSD score significantly predicted a higher help-seeking intent, except when including psychological violence in the model. The sensitivity analysis with the consistent reporting showed some additional significant associations of current mental diagnosis (negative) and severe physical illness (positive).

**Table 2.**
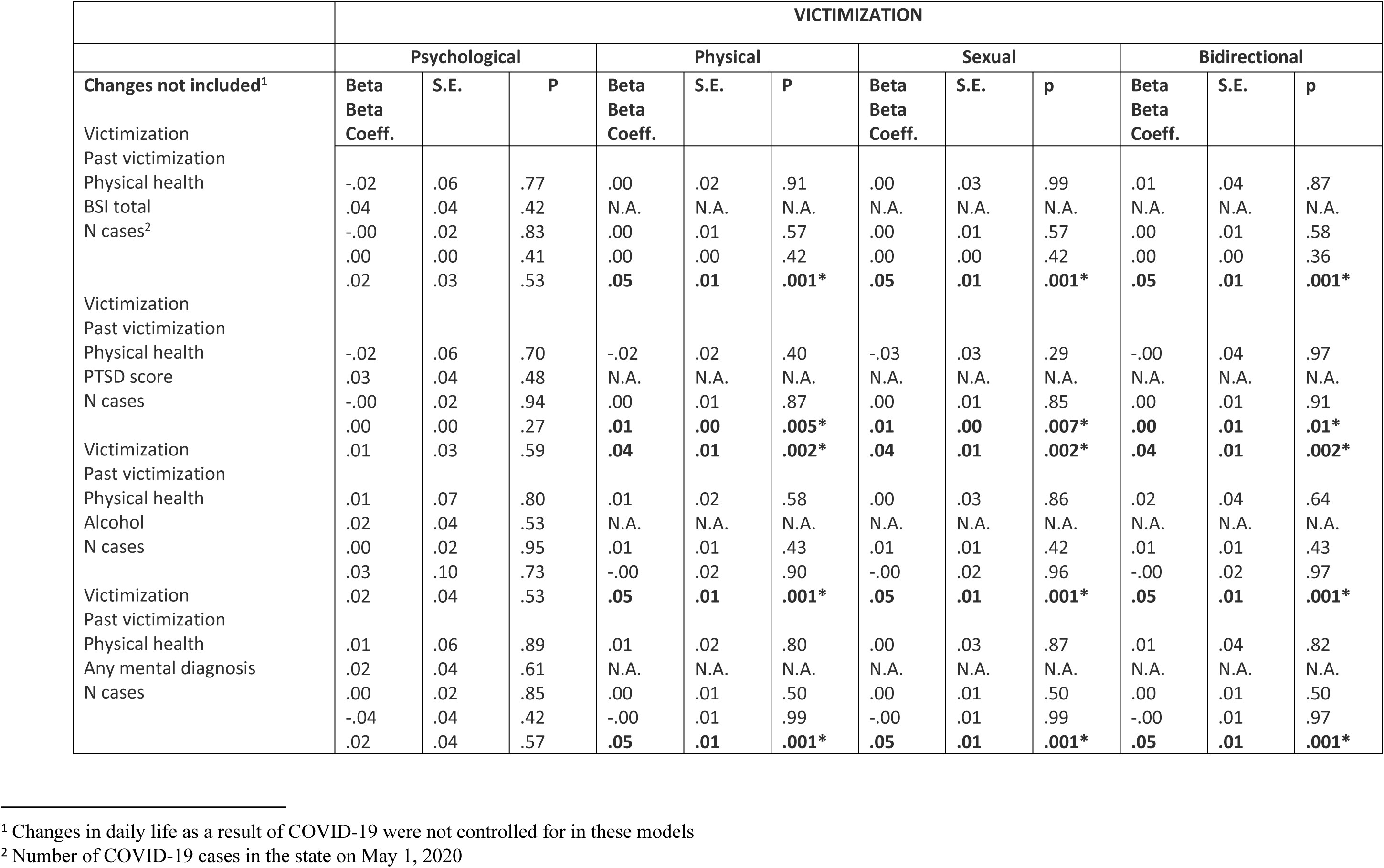
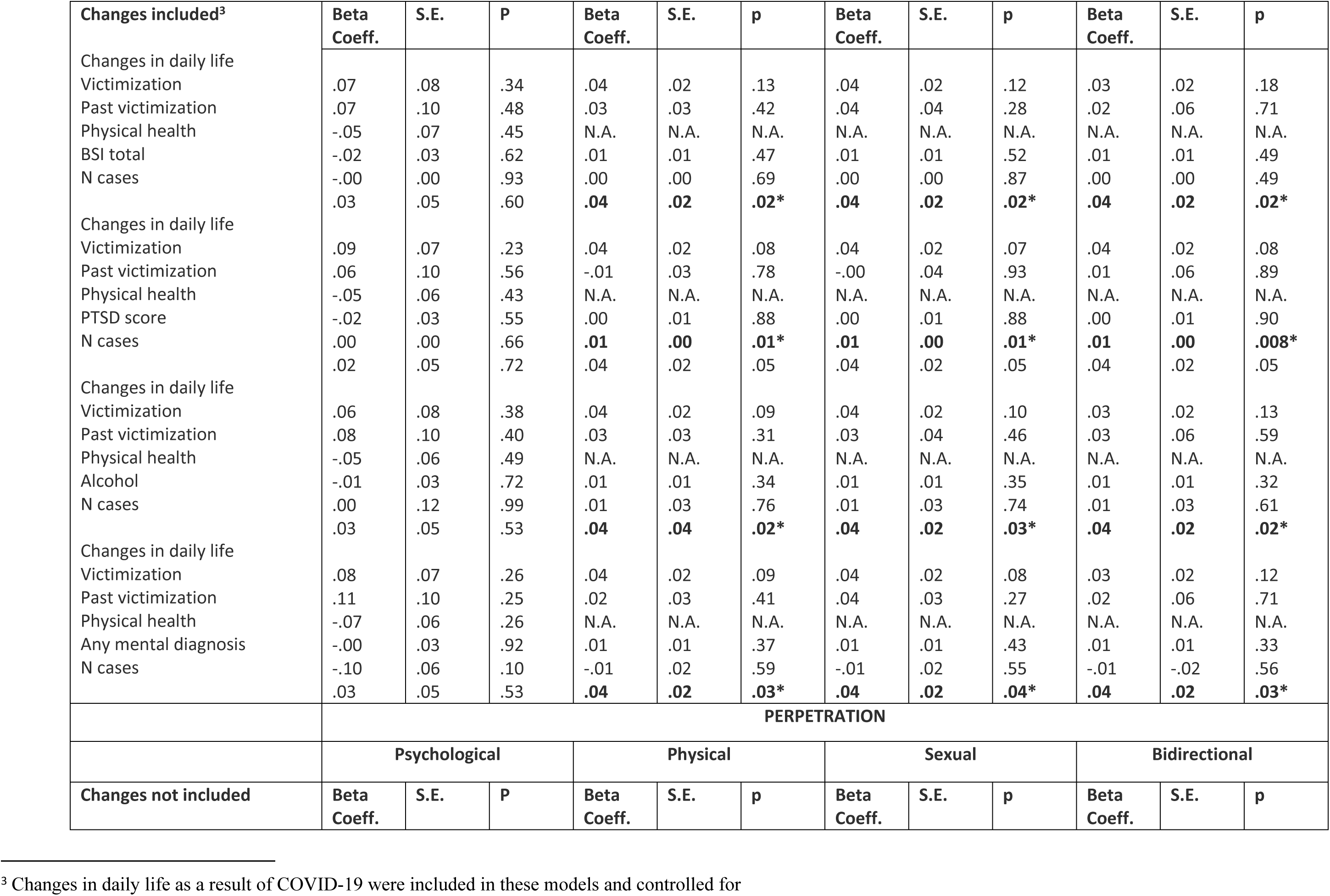
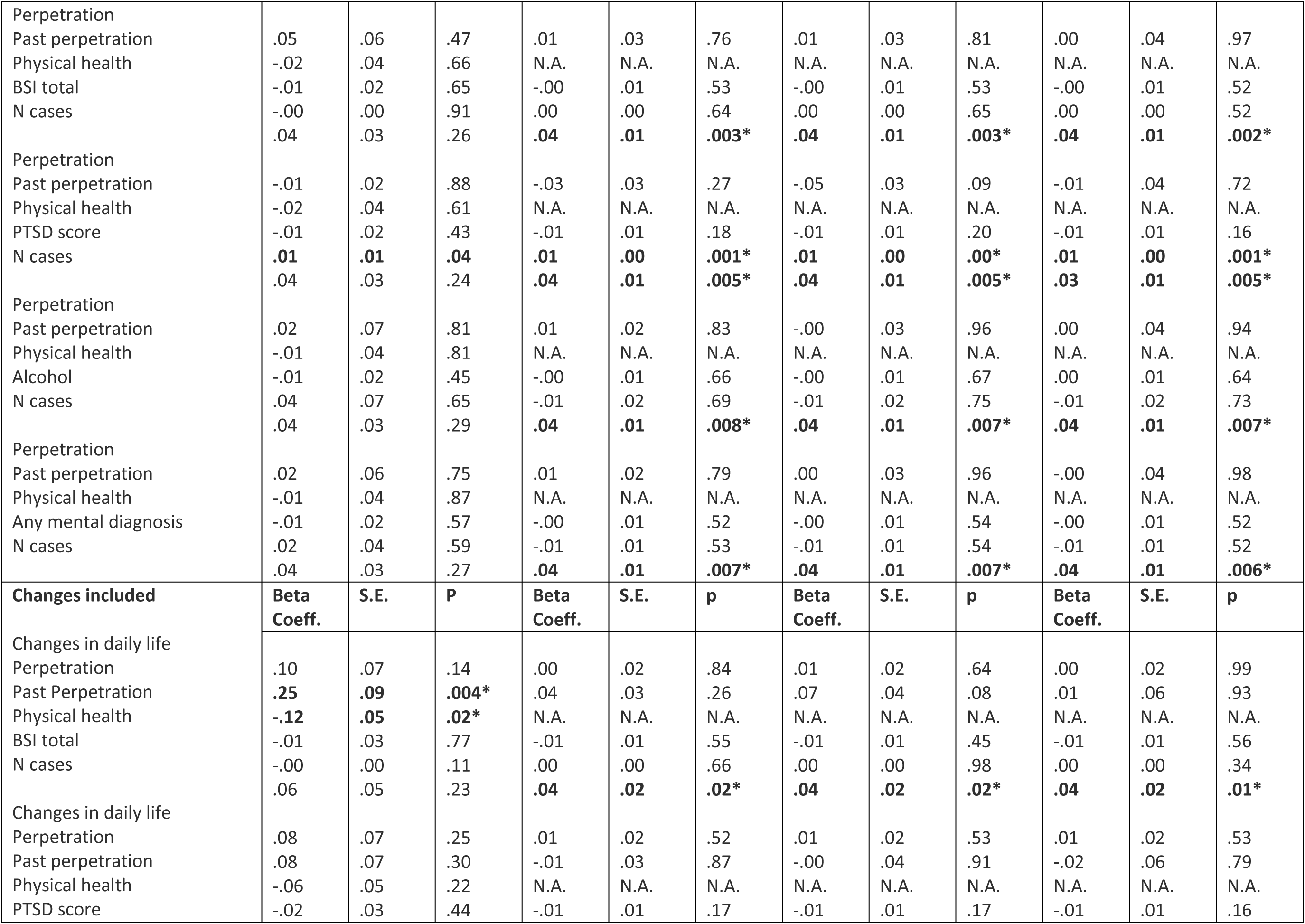
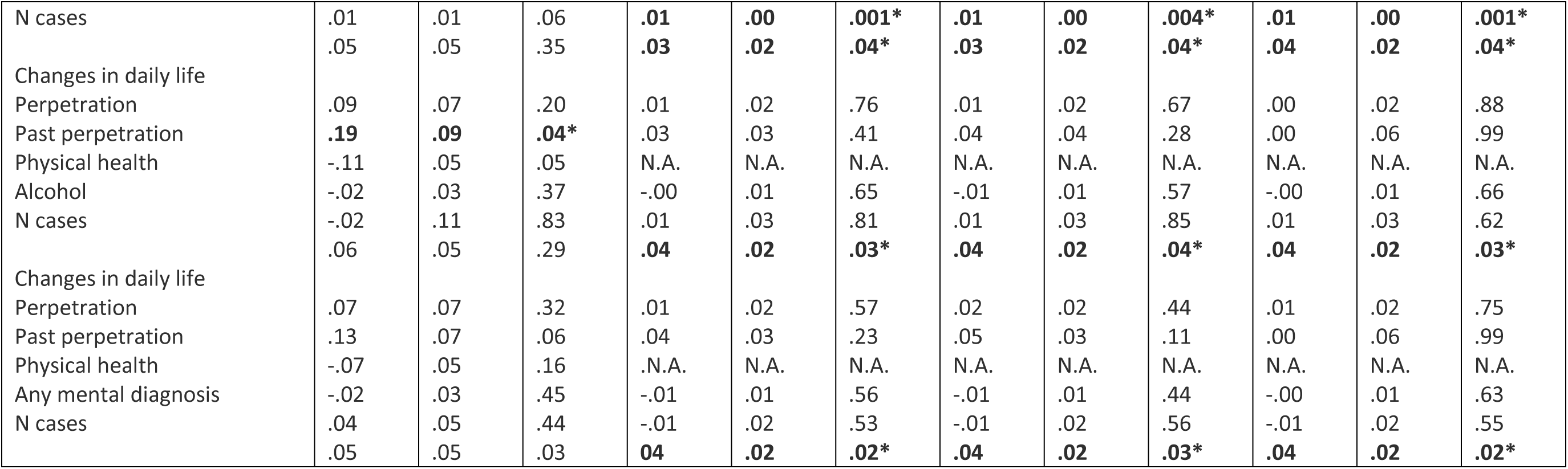
Help-seeking intent.

**Table 3.**
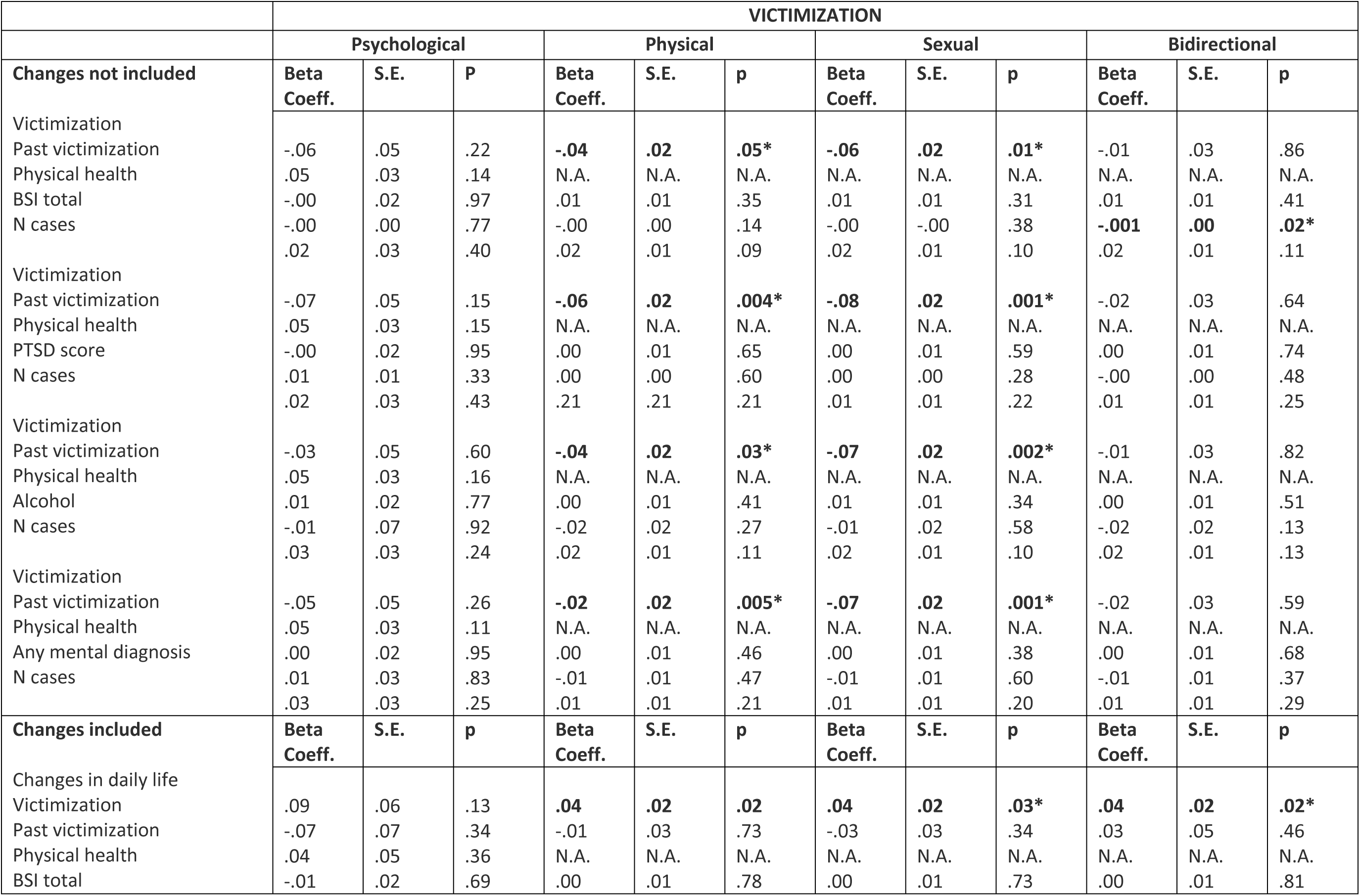
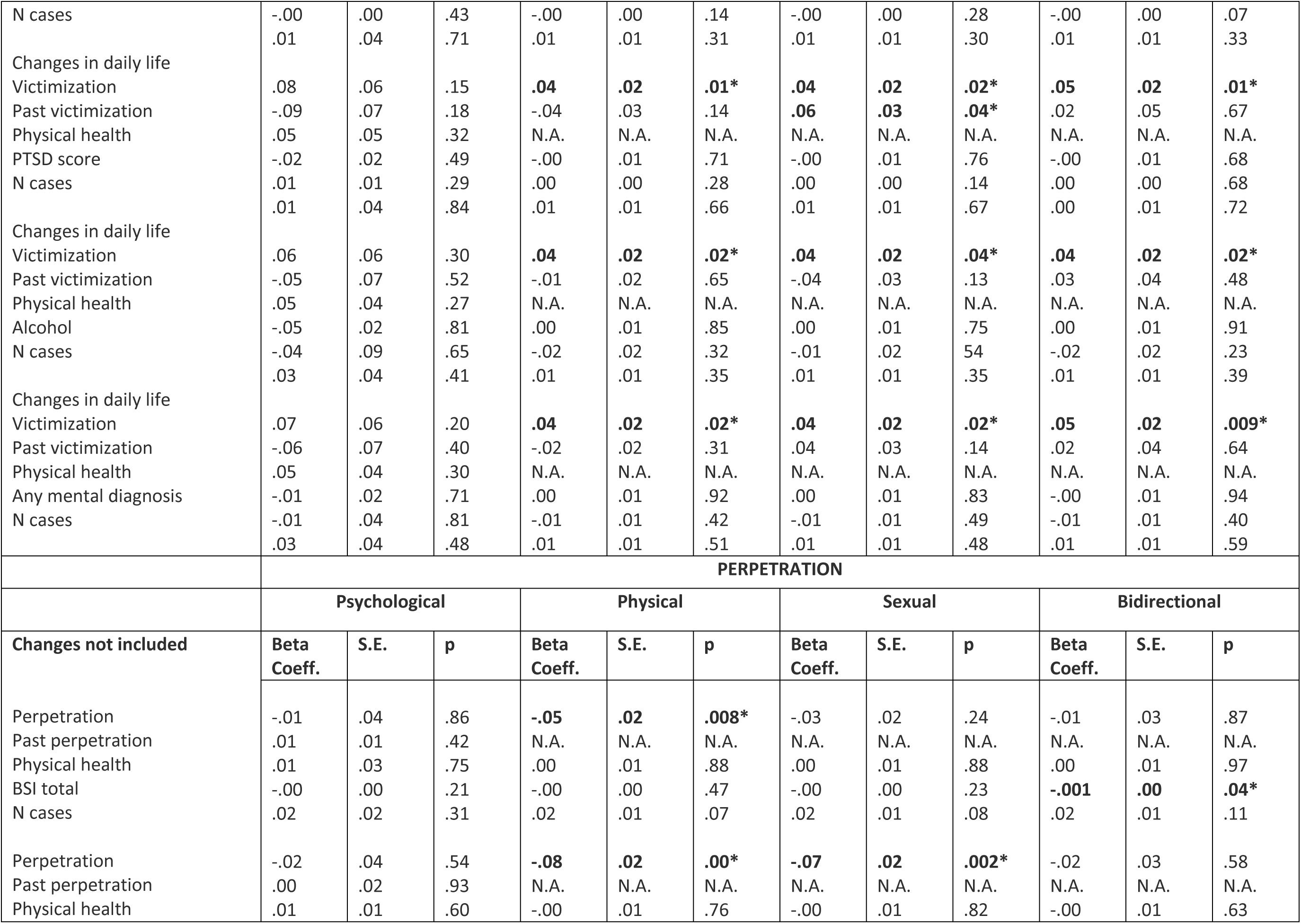
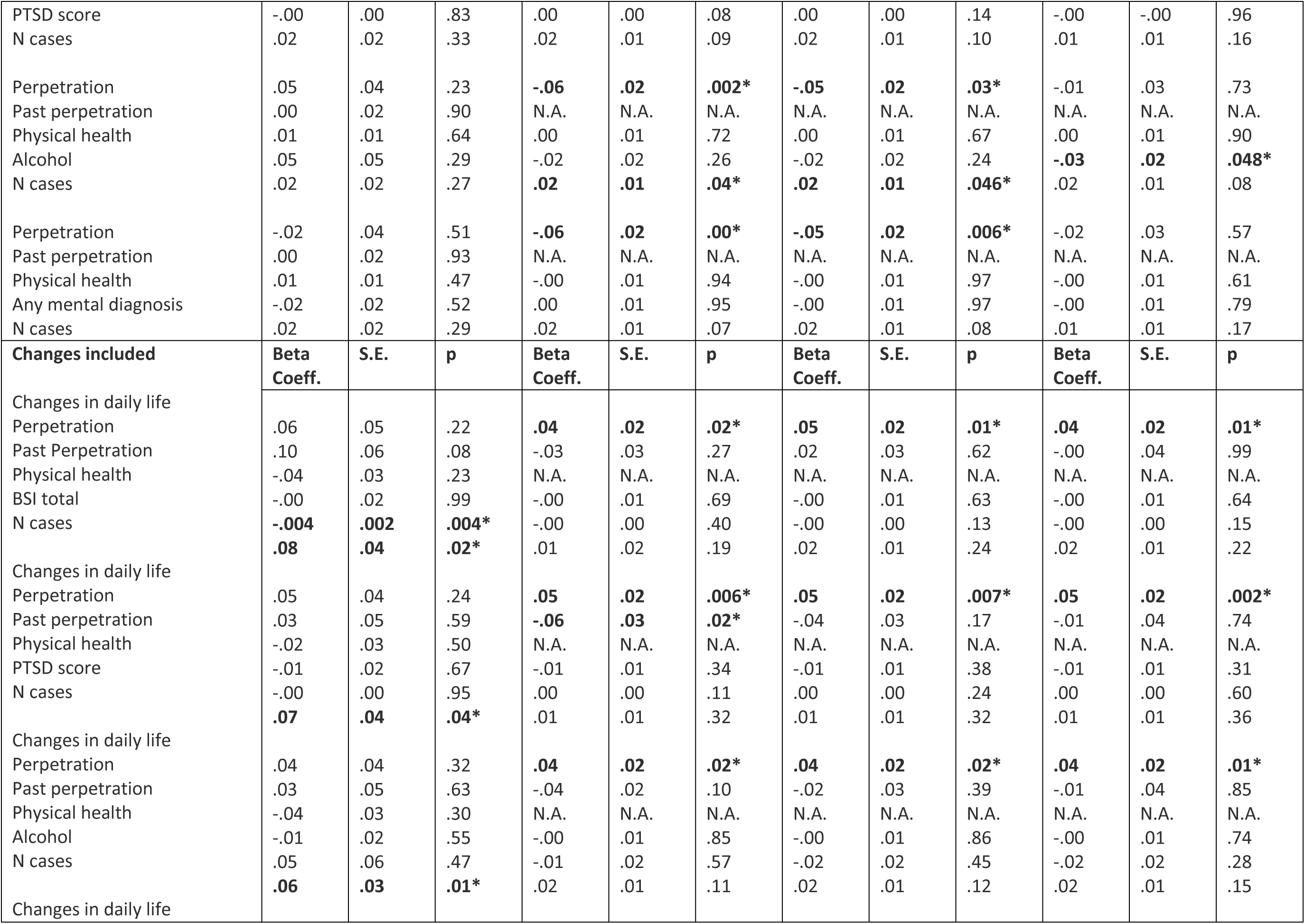
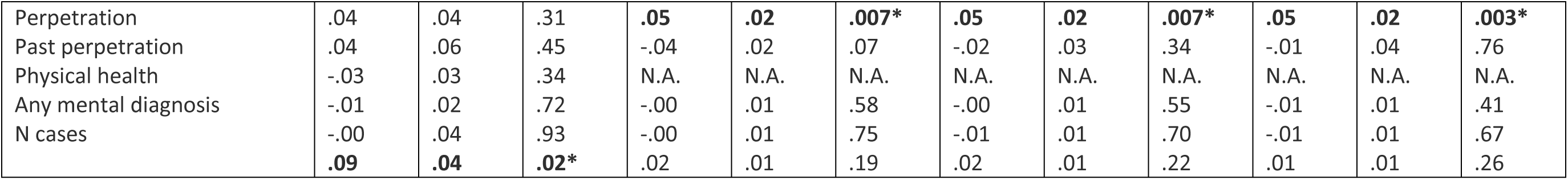
Disclosure intent.

#### RQ 1.2: Did correlates of help-seeking intention differ by gender? (Table 4)

Kruskal-Wallis tests showed different help-seeking intent among male survivors, and among perpetrators identifying as “other”, but only in the main analysis. In the sensitivity analysis of consistent answers across IPV scales, female perpetrators showed a lower help-seeking intent. The subgroup analysis among males gave the exact same results as the main analysis. No significant correlates were found for females in the main analysis; only in the sensitivity analysis with consistent answers across IPV scales, PTSD was a positive correlate and additionally among survivors current alcohol abuse was a positive correlate, and having a life-threatening illness a negative one, in contrast with the full sample. The participants identifying as “other” were fewer than 40 and most models could not converge or collinearity was too high.

**Table 4.**
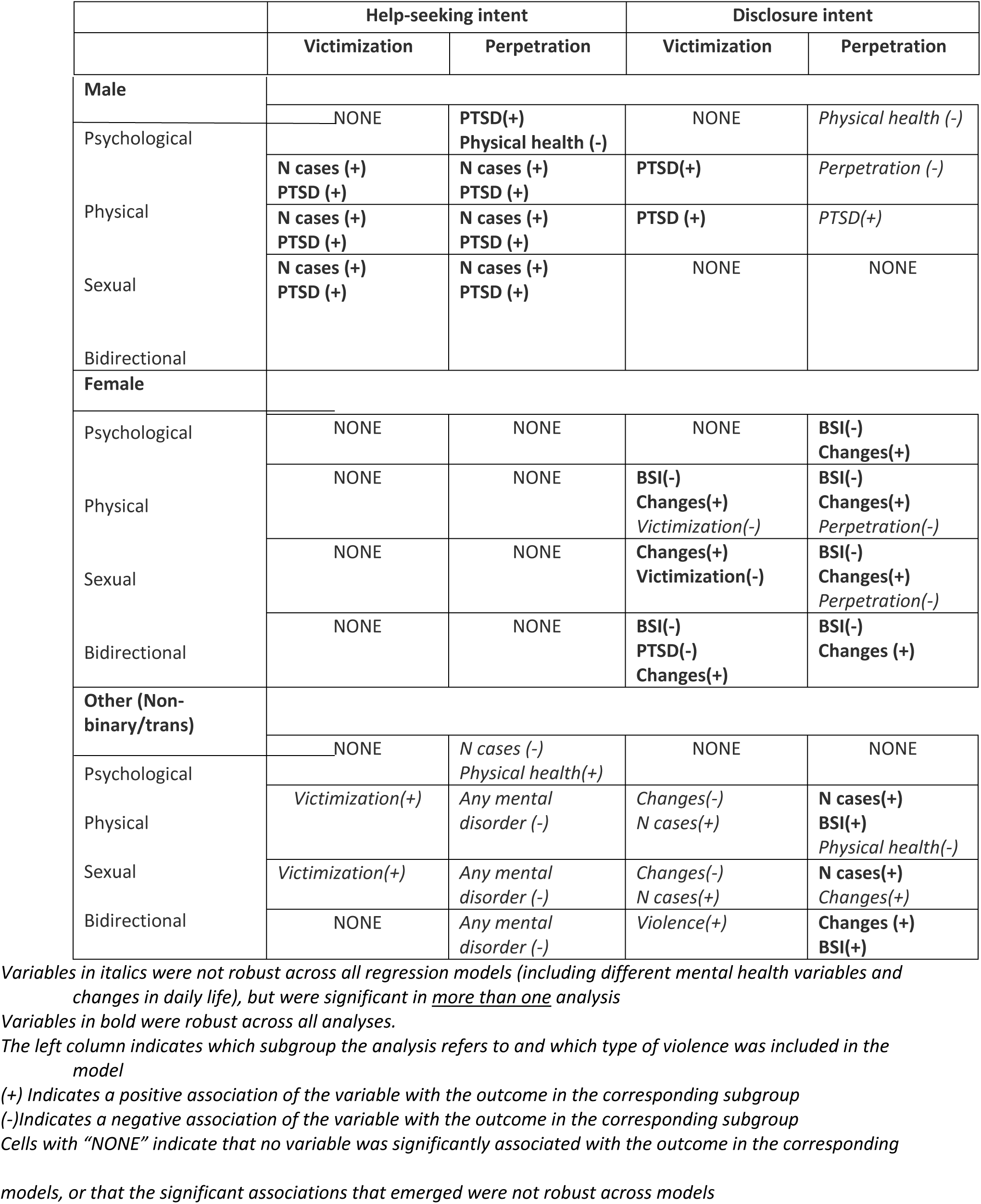
Significant variables in subgroup analyses by gender.

#### RQ 1.3: Did correlates of help-seeking intention differ by age? (Table 5)

Regarding age groups, adults had significantly higher intent to seek help than emerging adults and middle aged in the main analysis, but in the sensitivity analysis the only difference was that the middle-aged perpetrators had a lower intent than adults. The different analyses of emerging adults gave very different results, with no correlate being consistent. Among adults, the results were quite similar to the main analysis (PTSD and number of cases in the state positive correlates), but past or present severe physical health issues were a positive correlate in conjunction with psychological victimization among survivors only in most analyses. PTSD and current psychological victimization were positive correlates among the middle-aged as well. Similarly to the complete sample and the adults, the sensitivity analysis of consistent cases showed a positive correlation of severe and life-threatening illness, albeit only among perpetrators, and the negative correlation of current mental disorder only among survivors, along with a positive correlation of current alcohol abuse. The older adult participants were too few for most analyses.

**Table 5.**
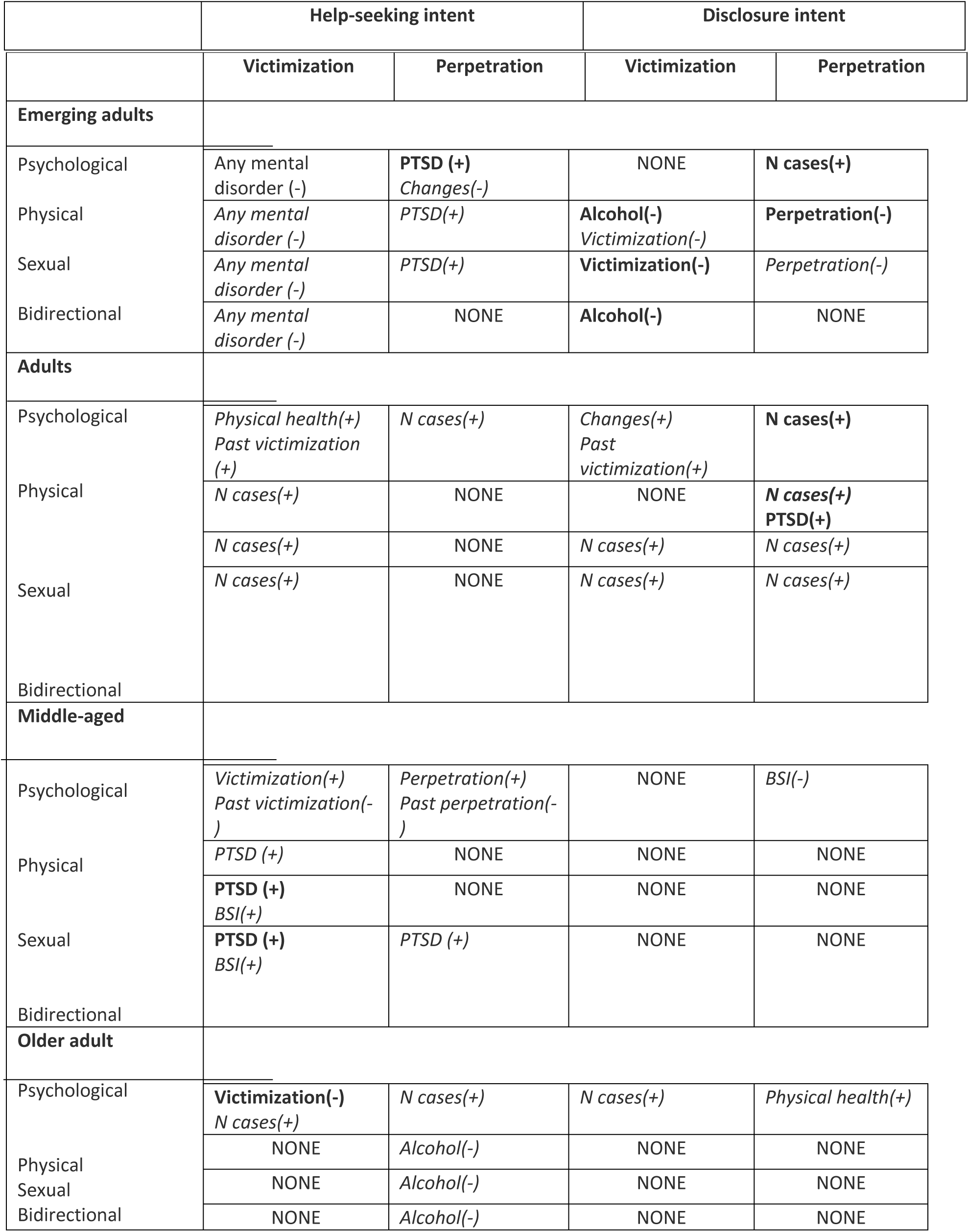
Significant variables in subgroup analyses by age.

#### RQ 1.4: Did correlates of help-seeking intention differ by income level? (Table 6)

Kruskal-Wallis tests detected that the two lower income groups were different from the two higher groups. Participants with low-income factors reached significance only when controlling for changes in daily life. These were a negative correlation of past psychological violence, but a positive correlation of current psychological violence. Bidirectional violence was negatively correlated. More factors reached significance in the sensitivity analyses, such as number of cases in the state (positive), and current mental diagnosis (negative), similarly to the full sample. Physical health issues, especially life threatening, but also mild ones, had strong negative correlations in the sensitive analyses, primarily among survivors, in contrast with the full sample. Among those with a middle income, number of cases was a significant correlate in many analyses. In the upper-middle income group, PTSD was positively associated. There were too few high income participants for analyses across all models.

**Table 6.** Significant variables in subgroup analyses by income level.

|  | Help-seeking intent |  | Disclosure intent |  |
| --- | --- | --- | --- | --- |
|  | Victimization | Perpetration | Victimization | Perpetration |
| **Low income** |  |  |  |  |
| Psychological | NONE | *Perpetration(+)*<br>*Past perpetration(-)* | NONE | NONE |
| Physical | NONE | NONE | NONE | NONE |
| Sexual | NONE | NONE | *Victimization(+)* | NONE |
| Bidirectional | NONE | *Violence(-)* | NONE | NONE |
| **Middle income** |  |  |  |  |
| Psychological | **Changes (+)** | *Changes (+)* | **Changes (+)**<br>**BSI(-)**<br>**PTSD(-)** | **PTSD(-)**<br>*BSI(-)* |
| Physical | **N cases (+)** | *N cases (+)* | **Changes (+)**<br>**BSI(-)**<br>**PTSD(-)**<br>*Victimization(-)* | **PTSD(-)**<br>**BSI(-)**<br>**Changes (+)** |
| Sexual | **N cases (+)** | *N cases (+)* | **Changes (+)**<br>**BSI(-)**<br>**PTSD(-)**<br>*Victimization(-)* | **BSI(-)**<br>**Changes (+)**<br>*PTSD(-)* |
| Bidirectional | **N cases (+)** | *N cases (+)* | **Changes (+)**<br>**BSI(-)**<br>**PTSD(-)** | **PTSD(-)**<br>**BSI(-)**<br>Changes (+) |
| **Upper middle income** |  |  |  |  |
| Psychological | NONE | **Physical health(-)**<br>*PTSD(+)* | NONE | *Changes(-)*<br>*PTSD(+)*<br>*Alcohol(+)*<br>*Any mental disorder(-)* |
| Physical | NONE | **PTSD(+)** | NONE | *Perpetration (-)* |
| Sexual | **PTSD(+)** | **PTSD(+)** | NONE | **PTSD(+)**<br>*Perpetration (-)* |
| Bidirectional | *PTSD(+)* | **PTSD(+)** | NONE | NONE |
| **High income** |  |  |  |  |
| Psychological | NONE | NONE | NONE | NONE |
| Physical | **PTSD(+)** | NONE | **PTSD(+)** | **PTSD(+)**<br>*N cases(+)* |
| Sexual | **PTSD(+)** | NONE | **PTSD(+)** | **PTSD(+)**<br>*N cases(+)* |
| Bidirectional | PTSD(+)<br>*BSI(+)* | PTSD(+) | PTSD(+) | PTSD(+)<br>*N cases(+)* |

#### RQ 1.5: Did correlates of help-seeking intent differ by ethnicity? (Table 7)

There were hardly any differences across ethnicities in help-seeking intent. Regarding correlates, the results of the White participants group were very close to the main analysis, with number of cases and PTSD being the most robust correlates, and low BSI score negatively associated with help-seeking among perpetrators of physical violence. No significant correlates were found for the Black participants group in the main analysis, but in the sensitivity analysis, when changes in daily life were included, they had a positive correlation. Among the Latinx participants, having a severe health issue was a negative correlate, in contrast with the full sample, and among survivors only having a mental disorder currently had a positive correlation. In contrast, among participants of other ethnicities, current mental disorders were negatively associated, and positive correlates were PTSD symptoms, severe physical disease, in line with the complete sample, as well as past physical violence.

**Table 7.**
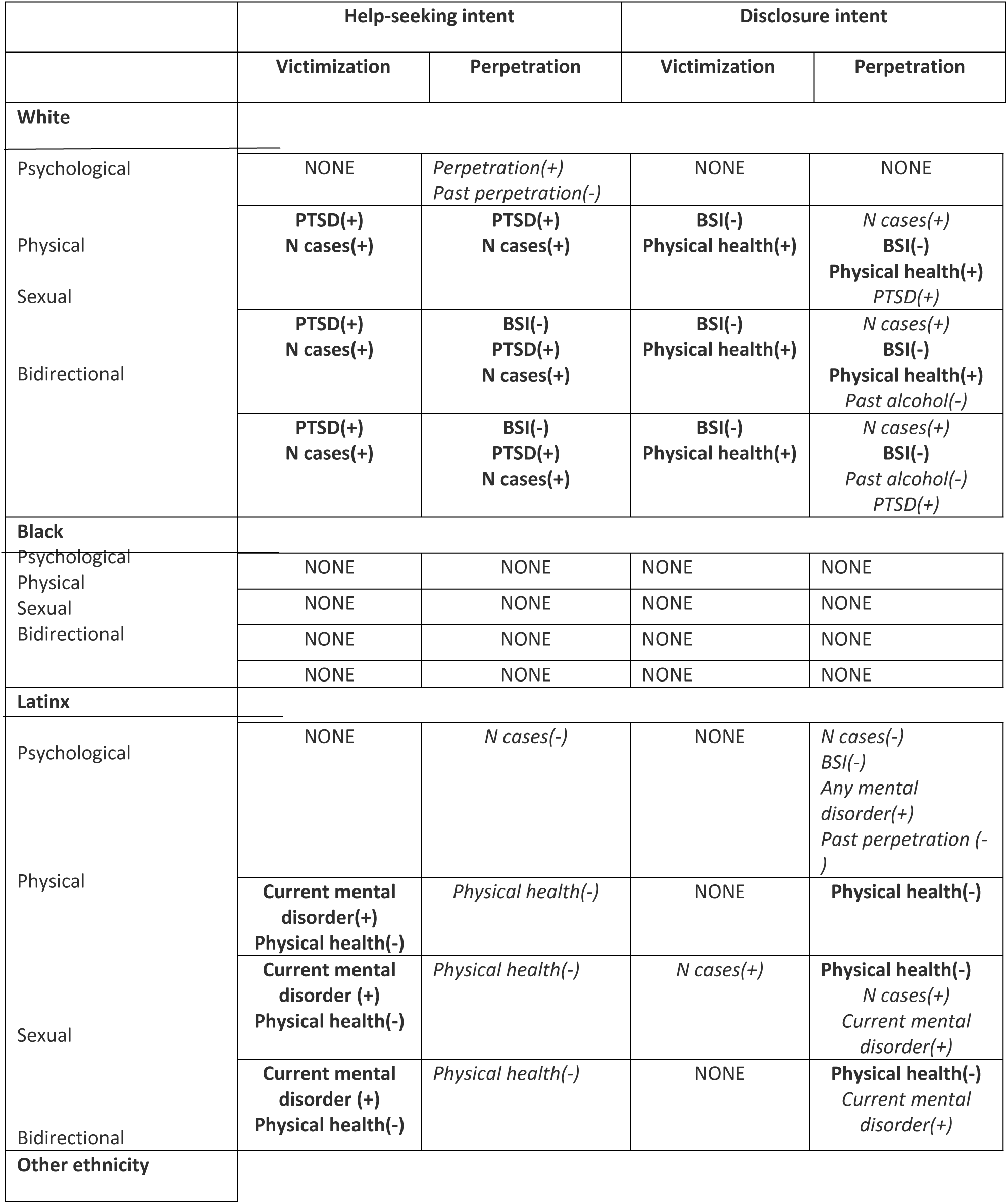
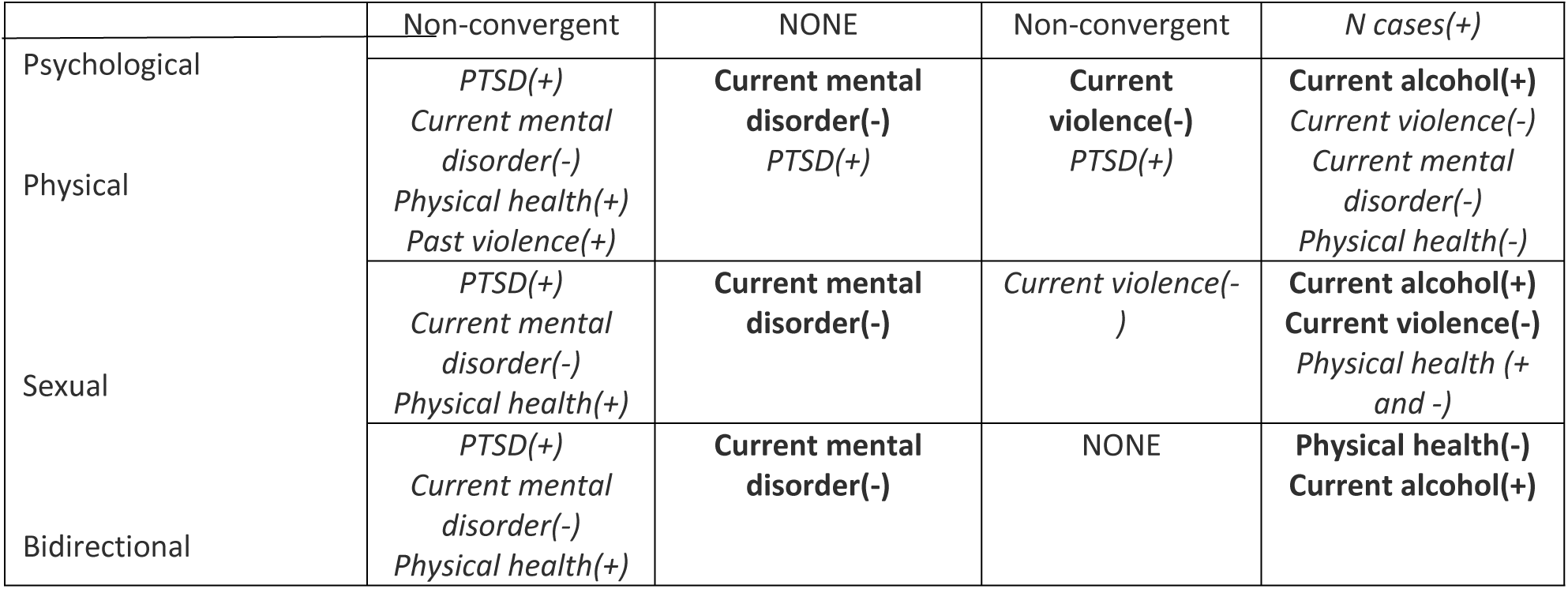
Significant variables in subgroup analyses by ethnicity.

### Disclosure intent

#### RQ 2.1: What predicted intention to disclose IPV? (*Table 3*)

Current sexual victimization was consistently negatively correlated with disclosure intent. BSI score was also negatively correlated, but less consistently.

#### RQ 2.2: Did correlates of intention to disclose IPV differ by gender? (Table 4)

Only the gender minorities group (i.e. “other”), reported significantly lower disclosure intent when using the E-HITS. J-IPV showed a higher disclosure intent of women. Among males, PTSD was often correlated with higher disclosure intent consistently, except when including psychological violence. Physical and sexual violence were negative correlates. This held among females, especially victims. Changes in daily life had some positive association among female victims and mixed associations of all physical health categories and alcohol use emerged in both sensitivity analyses. For gender minorities, there were too few participants to support all the models, especially the sensitivity analysis of consistent reporting.

#### RQ 2.3: Did correlates of disclosure intention differ by age (life-stage)? (Table 5)

Emerging adults showed overall significantly lower disclosure intent. The J-IPV also showed a lower disclosure intent amongst adults compared to older adults. Older adult survivors were significantly different from adults and middle aged. In emerging adults, similarly to the main analysis sexual victimization, and additionally alcohol abuse, any mental disorder and physical perpetration correlated with lower disclosure intent among survivors. In adults, number of cases had an overall positive association. More correlates became significant in the sensitivity analyses, which were compatible with the full sample, i.e. negative correlations of sexual and physical violence and BSI score. Among the middle aged, there were no consistently significant correlates in the main analysis. However, the sensitivity analyses yielded several significant factors. Current mental disorders had consistent negative correlations and PTSD consistently positive correlations. Among perpetrators, life-threatening illness had positive correlations. Among survivors, BSI had negative correlations and current alcohol abuse had positive associations. Among the older adults, the number of cases showed positive correlations and severe diseases had some mixed correlations.

#### RQ 2.4: Did correlates of disclosure intention differ by income level? (Table 6)

There were significant differences between income level groups, generally lower income groups had lower disclosure intent. In the low income group, only sexual victimization showed some consistency in the correlation with lower disclosure intent, similarly to the full sample. In the sensitivity analyses current illness of any severity and current physical perpetration showed negative correlations. Among those with middle income, no correlate was very robust. In the upper middle income group, sexual violence was a robust negative correlate in line with the full sample, and PTSD was a less consistent positive correlate. PTSD had a consistent positive association among those with high income, as well as number of cases with a positive correlation among perpetrators. In sensitivity analyses past alcohol abuse was negatively associated too.

#### RQ 2.5: Did correlates of disclosure intention differ by ethnicity? (Table 7)

When using the E-HITS in the main analysis, Black participants survivors and those of “other” ethnicity showed lower disclosure intent than the White participants and multi-ethnic. The same held for perpetrators, with the addition that Latinx participants perpetrators showed lower intent than the White participants and Native participants. These findings were not confirmed in the sensitivity analysis with the consistent cases. In terms of correlates, in the White participants group, BSI score had a negative correlation, as in the full sample, and severe physical illness had a positive association. The main analysis gave no significant correlates for Black participants, but the sensitivity analysis of consistent cases showed negative associations of different levels of the physical health variable, mixed association of physical violence, and some positive correlations of changes in daily life and BSI score for perpetrators, unlike the full sample. Among the Latinx participants, in the main analysis life threatening diseases had a robust negative correlation among perpetrators, but among survivors significant correlates were found only in the sensitivity analysis of consistent cases. These were the negative correlation of life-threatening diseases, as among the perpetrators, and past alcohol abuse had an ambiguous correlation. In the main analysis of survivors of other ethnicities current physical victimization was a robustly significant negative correlate, and less robust ones were sexual victimization and PTSD. Among perpetrators life-threatening disease was a robust negative correlate, and current alcohol abuse a positive one.

## DISCUSSION

The findings in summary showed that PTSD was the most robust correlate of all outcomes, especially help-seeking intent, and it had a positive direction in most cases. PTSD symptoms had a varying correlation across population groups on disclosure, which was more positive for men and people with a high income. The number of cases in the state also had a high impact on help-seeking intent, at least among males, people of White ethnicity, age 30-44 and lower income (especially for help-seeking intent), but often it was explained by the changes in daily life that the pandemic instigated. However, changes in daily life had some positive association, especially for women, Black participants and people of middle income. Disclosure was impacted negatively by physical and sexual violence. Alcohol abuse had mixed to negative correlations, especially among emerging adults and women. Inconsistencies in self-reported violence across different scales significantly impacted results, leading to more significant findings when only participants with consistent reports were analyzed. This issue is highlighted particularly among emerging adults, middle aged, people of middle and upper middle income, Black participants and Latinx participants, and perpetrators. The lower help-seeking and disclosure intent of gender minorities and emerging adults highlight their vulnerability and a need for additional attention.

All in all, external factors, in this case the number of COVID-19 cases in the state, impacted more help-seeking intent, especially in the analysis of all participants regardless of consistency in reporting. There are highly considerable nuances across populations. Changes in daily life are a sort of an in-between factor for the external and intra-individual effects, and they appear to confound the associations of number of cases.

The positive impact of number of COVID-19 cases on help-seeking was unexpected, and it diverges from the literature showing decreased contacts with healthcare services during the first and strict stages of the lockdown (42, 76, 77). It is probably explained by the fact that this study measured intention, instead of actual behavior; most importantly, the phrasing of the questions “Were a healthcare provider to ask you such questions…” refers to a positive scenario where a healthcare provider offers resources, and it sounds as if the availability of these resources is not questioned, and this barrier could thus be considered as skipped. In such a case, a provider offering resources might come as a positive surprise eagerly accepted. Alternatively, it may also be a sense of urgency that could have been created in states overburdened with COVID-19.

PTSD symptoms had mostly positive associations, which is in line with the previous findings of mental ill health as motivation to accessing care (40, 78, 79), and especially the high association of PTSD with service use (73, 80). The mixed correlations of the BSI score are also in line with the literature and the theory that support a varied role of mental health (39, 41). The results on negative impact of violence initially appear incongruent with the evidence of severity of violence being highly correlated with disclosure and help-seeking (78) (81, 82) and also from some evidence of increased disclosure during the pandemic by survivors of coercive control (83), but the present study did not assess the severity of physical and sexual violence, or the exact concept of coercive control, so it remains unclear. It might also be due to the differences between intent and actual behavior. The negative correlations of mental health related variables that were detected could be interpreted as pointing to the fear and stigma that are frequently found in the literature as barriers to disclosure (16, 17, 84–86). Maybe the PTSD symptoms facilitate the recognition of mental health related struggle and are thus contributing to help-seeking.

One of the least previously investigated variables and a key aspect of the pandemic context, was changes in daily life. Interestingly, it accounted for a considerable part of the variance, perhaps because the mental health-related and violence variables are connected to these changes in daily life, and perhaps are confounding. Their positive correlations with disclosure intent might be related to a sense of urgency the pandemic created, and may be related to additional pressure.

Another noteworthy finding was that the correlations were often similar for survivors and perpetrators, but this could be attributed to the high prevalence of bidirectional violence in the sample. This fact might also account for the underscored role of PTSD symptoms in this study, as PTSD is a very important factor in bidirectional violence (87).

An element that stands out is the considerable differences across sensitivity analyses, which highlights the fact that many people do not report on violence consistently across scales. This could be a sign of hesitation to disclose or maybe uncertainty as to whether their experiences qualify as abuse. The phrasing of the questions appears to matter, which is not surprising given how challenging it can be to talk about abuse. This might mean that it would be even more difficult in a healthcare service, especially in the context of the pandemic, compared to the anonymity of an online survey. The fact that a lot more factors reached significance in the sample with consistent answers across IPV scales could be interpreted in many ways; possibly the individuals reporting consistently share more similarities and are more affected by the factors such as mental and physical health. Emerging adults, Black participants and Latinx participants’ pronounced inconsistency is likely indicative of a vulnerability by being reluctant to disclose either because of doubts or fear of the consequences. The primarily negative correlations of physical illness for Black participants and Latinx participants are possibly linked with unfavorable medical insurance, as a structural barrier to help-seeking (88). The underlined inconsistency of the higher income groups is harder to interpret.

Regarding the evidence produced by the subgroup analyses, the lack of impact of many of the independent variables on help-seeking intent among women in the main analysis, could be interpreted as a failure to investigate factors that matter more to women, such as having to care for children (89, 90), or financial issues and dependence from their partner. There were also no significant correlates of disclosure among the Black participants and the Latinx participants in the main analysis, so more determining factors, possibly immigration status or trust and access to the services. It was also challenging to detect significant correlates for the low income group, despite having sufficient participants, which could also reflect that factors not investigated in this study, such as financial dependency on the partner, which could be more determining (19, 86).

Men in this sample appear to be more empowered to disclose IPV victimization than described in the literature, where men have been found to face elevated stigma (72). This may be because the pandemic and its socio-economic consequences had an overall lower burden on men (91). People of age 30-44, also seem to be quite empowered and relatively more open to seeking help, and the same holds for the higher income groups, who appear less influenced by external factors and violence, and where mental health issues serve as motivation to open up to options. On the other end, emerging adults (18–29) seem to be less open to seeking help and disclosing, which corroborates the findings of some of the previous studies (71), but is at odds with others (28), presumably because it is a group more negatively affected by poor mental health, especially survivors. The inconsistencies in reporting of the Black participants and the Latinx participants could be seen as a vulnerability. Overall, the results of the subgroup analyses point towards a sharpening of pre-existing economic and racial vulnerabilities and inequalities, which is congruent with qualitative research among Black participants survivors (92) and minorities in general in high income countries (93, 94).

These findings should be interpreted in the light of the strengths and limitations of this study. The major strength is that the individuals were reached directly, instead of inferring the barriers to help-seeking just by the service use records. However, the limitations are not to be overlooked. First and foremost, the phrasing of the outcome questions, especially on help-seeking intent, skip the reality of limited services and touch more upon an ideal scenario. Secondly, the cross-sectional design allows only for associations and does not elucidate the temporal order of the factor and the outcomes. Thirdly, the statistical analysis chosen cannot explain the interplay among variables, neither the factors tested, nor the demographics that would provide evidence on intersectionality. Another important source of doubt is the multiple models tested, which increases the chance of Type I error and some variables reaching significance due to chance. For that reason we tried to report and emphasize mostly the results that were replicated across models.

To overcome the pitfalls of this study, future research should use longitudinal designs when possible in order to establish the temporal order, phrase the questions more directly, inquire about actual behavior as well, in order to examine to that extent intention translates into behavior. Structural equation modelling would also be a useful technique, as it would allow for investigating more complex relationships among the variables. Factors influencing help-seeking among women and people of color in the context of the pandemic warrant further investigation. Further research on intersectionality is also necessary, especially given that the results imply that pre-existing inequalities, at least those related to income level and ethnicity, persist in the context of the pandemic, and qualitative studies provide evidence towards that direction (8). Moreover, it was not possible to thoroughly investigate the help-seeking and disclosure intent for gender and sexual minorities, but there is an indication that their disclosure intent is lower, so it would be important to investigate that further.

Despite these limitations, valuable practical recommendations remain. Most importantly, the approach of the clinicians should be as careful and encouraging as possible, as the phrasing does matter and many people are not very willing to report IPV. Plus, it is important to screen in the presence of mental distress even if there are no signs of physical abuse (95), and also because it is already evident that provision of online services in the pandemic context ought to look out for survivors, instead of waiting for them to reach out (96). There is an indication that people aged 18-29, and those of lower income face more barriers to help-seeking, and they should be prioritized. Emerging adults, the highest risk age group for IPV (97), should be prioritized. Men appear to be comparably involved in IPV, both as perpetrators and survivors, so providers should not neglect them, but instead *abolish stereotypes* towards them. Screening that builds upon trauma symptoms while remaining mindful of overall distress symptoms has a higher chance of being successful, as traumatic stress symptoms have a strong positive association with help-seeking and disclosure intent for many demographic groups, but overall distress and mental disorders have more negative correlations. It would also be helpful if mental health services focused more on screening for IPV, especially because mental health appears so closely intertwined with IPV. Given the rise of online mental health interventions, integrating IPV screening and service linkage is crucial, as survivors—especially vulnerable groups—report willingness to engage online (98, 99). Online screening seems more feasible in this context (100), but also been shown to be more acceptable and effective (101). This should be implemented by taking serious privacy precautions, given that concerns on safety and privacy issues of online services have been raised in the context of the pandemic (102) (10). However, it is important that some in-person services continue flexibly through collaboration of different services and conscious proactivity of providers (103), not to heighten the digital inequalities (96).

## CONCLUSION

The context of the COVID-19 pandemic was a challenge for IPV involved people and healthcare providers, but when seen as an incentive for change, there is potential. Careful and encouraging approach by healthcare providers is highly recommended in the event of emergency situations, such as natural disasters or potential future health crises. Even more attention should be paid to historically vulnerable groups, because the services and programs that are typically restricted in emergency contexts are those that serve these groups (104).

## Data Availability

The data supporting this study are available upon reasonable request from the second author, Dr Maxine Davis.

